# Restrictions on top speed and nighttime usage substantially decrease the incidence of electric scooter injuries

**DOI:** 10.1101/2023.06.20.23291641

**Authors:** Oskari Pakarinen, Arja Kobylin, Veli-Pekka Harjola, Maaret Castrén, Henri Vasara

## Abstract

**Objective:** To investigate the effectiveness of restrictions on top speed and nighttime usage on the incidence of e-scooter-related injuries.

**Design:** A retrospective comparative study. We compared shared e-scooter injuries from two periods: Unrestricted period (1.1-31.8.2021), and Restricted period (1.1.-31.8.2022).

**Setting:** General population of Helsinki, Finland. We collected the data from the electric patient database from three trauma hospitals representing all public hospitals treating acute trauma patients in Helsinki.

**Participants:** All patients with an injury related to shared e-scooter riding sustained in Helsinki

**Interventions:** The restrictions established for shared e-scooters during the restricted period were: 1) The daytime top speed of 20km/h, as opposed to the previous 25km/h, 2) the use of shared e-scooters was prohibited on Friday and Saturday nights between 12 p.m. and 5 a.m, and 3) the nighttime top speed was lowered to 15 km/h from Sunday to Thursday between 12 p.m. and 5 a.m., as opposed to 25 km/h.

**Main outcome measure:** Proportional incidence of e-scooter injuries compared to the total trips made by e-scooters.

**Results:** There were 528 e-scooter injuries requiring hospital care during the unrestricted period (1.1.-31.8.2021) and 318 injuries during the restricted period of similar length (1.1.-31.8.2022). The proportional incidence of e-scooter injuries was 19 (95% CI 17-20) for every 100 000 rides during the unrestricted period and 9 (95% 8-10) per 100 000 rides during the restricted period. In the risk analysis, the odds ratio (OR) for shared e-scooter accidents was 0.5 (95% CI 0.4-0.6) for the restricted period when adjusted for hourly temperature, rain amount, wind speed, and visibility. After introducing the restrictions, the number of e-scooter injuries decreased significantly between 11 p.m. and 5 a.m.

**Conclusions:** Restrictions on the top speed and nighttime usage of e-scooters decreased the amount of e-scooter injuries. We recommend similar restrictions in cities with shared e-scooter services.

## Background

Electric scooters (e-scooters) have become a popular means of transport in large cities worldwide. Rental companies providing shared e-scooters have played a major role in the increased use of these vehicles (1). The e-scooters have the potential to establish themselves as useful and environmental-friendly urban means of transport. However, in recent years, e-scooter-related accidents have become a serious health issue worldwide (2–4). As a result, for example, the city of Paris plans to ban e-scooters from September 2023 after a public vote where approximately 90% of participating citizens voted against using these vehicles.

Typical e-scooter injuries include head injuries, fractures of extremities, and superficial wounds (5–7), but life-threatening injuries have also been reported (5,8). The rate of e-scooter injuries increases in late hours (7,9), and a substantial part of the patients who suffer an injury with an e-scooter are driving intoxicated (8,9). Accordingly, the need for e-scooter regulations has been brought up in numerous instances (10,11).

In 2021, there were 446 e-scooter injuries in the city of Helsinki (12). To limit the injuries caused by e-scooters, the local government set regulations for shared e-scooter usage in co-operation with the e-scooter rental companies. These restrictions limited the top speed and nighttime use of e-scooters (13). The objective of this study was to investigate the effect of these restrictions on the incidence of e-scooter-related injuries. We hypothesised that the restrictions would decrease the incidence of injuries.

## Patients and methods

### Design

We conducted a retrospective comparative study at the Helsinki University Hospital, where we compared e-scooter injuries from two periods: 1) Unrestricted period (1.1-31.8.2021), and 2) Restricted period (1.1.-31.8.2022, with restrictions on e-scooter top speed and availability on nighttime). The restrictions were introduced at the beginning of September 2021, but we decided to exclude the end of 2021 to have an equal comparison period (from January to August) with and without restrictions. The data was collected from a collective electric patient information system from three trauma hospitals representing all public hospitals treating acute trauma patients in Helsinki: two level I trauma centres and one level IV trauma centre.

### Setting

The city of Helsinki has 656 920 residents, with a mean age of 41 years (14). Shared e-scooters have been available since 2019, and the availability of shared e-scooters has since been increasing. In 2021, there were four operators in action, with an average fleet size in the busiest operational season (May to August) being 6121 shared e-scooters. In comparison, in 2022, there were as many as six operators, with an average fleet size of 15612 e-scooters, respectively. Users over 18 years old can rent e-scooters with a mobile application. According to Finnish legislation, the maximum speed of e-scooters is 25 km/h. Wearing a helmet is strongly advocated but is not controlled publicly. Driving under the influence is forbidden by the law and the rules of shared e-scooter companies. However, there is no breath alcohol penalisation limit regarding e-scooters, thus, making effective surveillance inexecutable.

### The public intervention

As a countermeasure to the rising number of e-scooter accidents, the city of Helsinki and the e-scooter rental companies constituted restrictions affecting rental e-scooters in the Helsinki area. On the 3^rd^ of September 2021 following restrictions were established to decrease the incidence of injuries:

1. The daytime top speed was limited to 20 km/h as opposed to the previous 25km/h,
2. the use of rental e-scooters was prohibited on Friday and Saturday nights between 12 p.m. and 5 a.m., and
3. the nighttime top speed was lowered to 15 km/h from Sunday to Thursday nights between 12 p.m. and 5 a.m. (13).

### Outcome

The primary outcome was the proportional incidence of shared e-scooter injuries compared to the total rides made by e-scooters. In addition, we compared the proportion of intoxicated drivers, helmet usage, and the total amount, severity, and temporal aspects of the e-scooter accidents.

### Data collection and inclusion

The patients were identified from Helsinki University Hospital data pool utilising a word search from the emergency department (ED) patient records. We used six e-scooter-related words with their inflected forms to identify the patients.

The ED patient records were investigated manually by authors OP or HV. We included all patients with an injury related to shared e-scooter riding sustained in Helsinki. Patients injured by private e-scooters and pedestrians injured by parked e-scooters were not included in the analysis. If intel regarding e-scooter ownership was unavailable, the patients were included in the analysis. All e-scooter accidents sustained in another city were excluded regardless of being treated in the Helsinki University Hospital.

We recorded patient characteristics and other necessary information regarding the accident from the ED records. The time of injury was set as accurately as possible according to the ED text. Otherwise, the time of the injury was set as the time the emergency contact call began or the patient was admitted to the ED. The patient’s most severe injury was graded based on the Abbreviated Injury Score (AIS) from the Abbreviated Injury Scale (15). The New Injury Severity Score (NISS) was calculated to estimate the total effect of all injuries on the patient (16). The breath alcohol level was recorded if measured at the ED or ambulance. In addition, alcohol intoxication was assessed as a binominal value, also considering the clinical assessment of the ED doctors if the breath alcohol level was not measured.

The usage data from the e-scooters was given to us by the Helsinki transport committee from the service Vianova Cityscope (Vianova SAS, Tournus, France, 2022). The data was handed in following Vianova’s terms of service regarding municipality intellectual property. The anonymised data included the overall amount of trips, driven distance, and fleet size, reported with hourly accuracy.

We sought data from the weather variables from both study periods from the Finnish Meteorological Institute open data (17). We included measurements from the temperature (°C), visibility (m), rain (mm), and wind (m/s) reported with hourly accuracy from the Helsinki Kaisaniemi measurement facility.

### Statistics

We presented nominal values as counts (percentages) and continuous values as medians or means based on whether the values complied with Gaussian distribution. Medians were reported with interquartile range (IQR), and means were reported with standard deviation (S.D.). The normality of continuous values was assessed visually using histograms and Q-Q plots and with the skewness value of the distribution. 95% confidence intervals (CIs) for proportional injury incidences were calculated using the Wilson score interval.

Binary logistic regression on a trip-by-trip basis was used to calculate odds ratios (ORs) with 95% CIs. In addition, we used Fisher’s exact test to test the statistical significance when comparing secondary outcomes. The analyses were conducted using the statistical program SPSS 29.0.0.0 (IBM corp. released on the 17th of November, 2022). We set the level for statistical significance as 0.05.

## Results

Overall, there were 528 e-scooter injuries requiring hospital care during the unrestricted period (1.1.-31.8.2021) and 318 accidents during the restricted period of similar length (1.1.-31.8.2022), respectively (Table 1). In total, 73 patients who were injured in another city, 18 patients who were injured by private e-scooters, and 17 pedestrians who were injured by parked e-scooters, were excluded from the analysis.

**Table 1:**
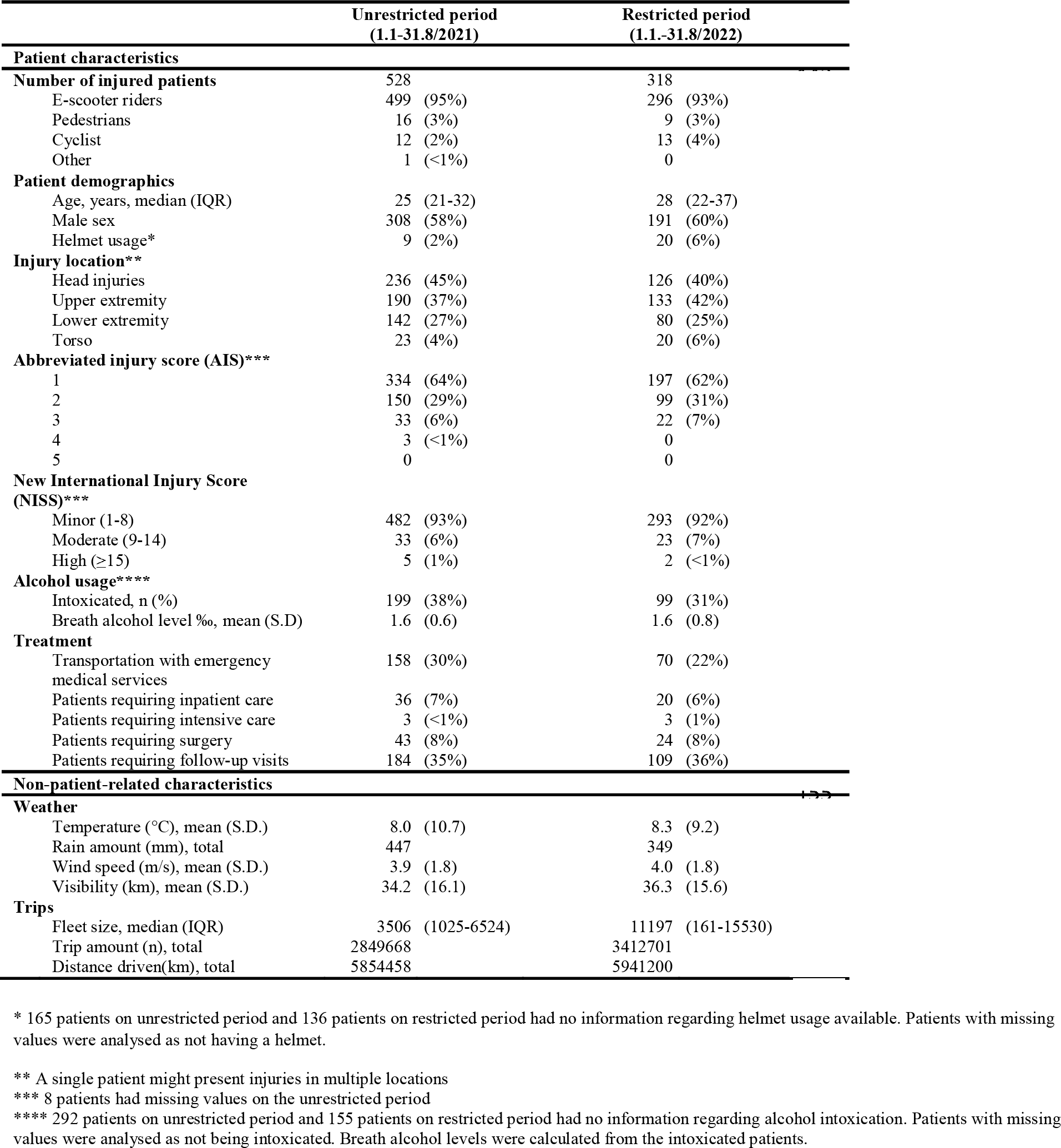
Characteristics of the study periods

The proportional incidence of all e-scooter accidents was 19 (95% CI 17-20) for every 100 000 rides during the unrestricted period and 9 (CI 8-10) per 100 000 rides during the restricted period when the top speed was lowered to 20km/h and nighttime restrictions were effective (Figure 1). Proportional to driven distance, the incidence was 9.0 (CI 8.3-9.8) injuries per 100 000km driven in the unrestricted period and 5.4 (CI 4.8-6.0) per 100 000km in the restricted period.

**Figure 1:**
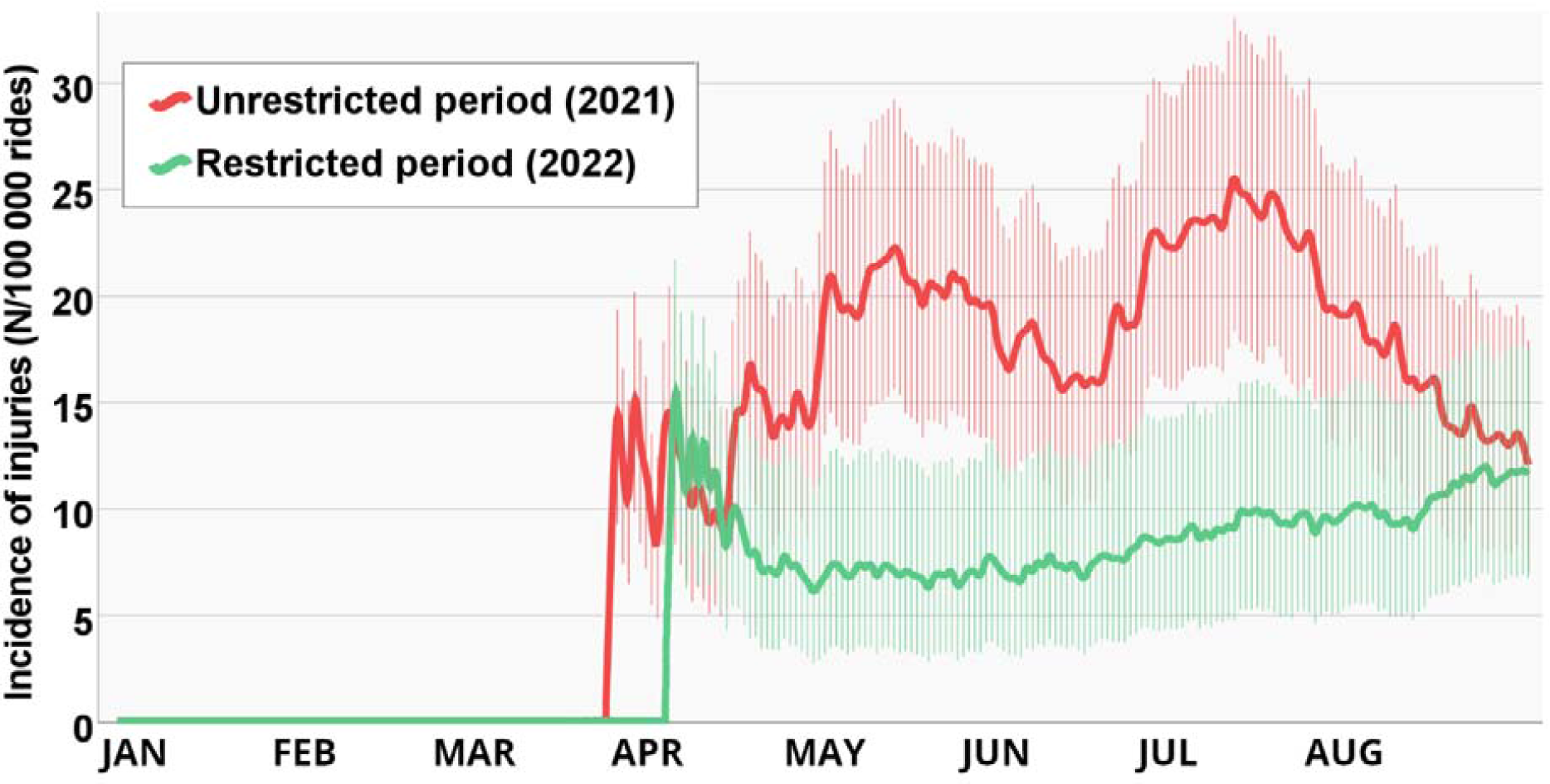
Incidence of e-scooter injuries per 100 000 rides. Each point represents the average incidence from the last 30 days. Injuries from periods with less than 10 000 rides in the last 30 days are not shown. The vertical lines indicate 95% confidence intervals. The restrictions afflicted: 1) Maximum speed limit 20km/h 2) Shared e-scooters were prohibited on Friday and Saturday nights between 12 p.m. and 5 a.m. 3) A nighttime speed limit of 15km/h for shared e-scooters between 12 p.m. and 5. a.m. from Sunday to Thursday The differences from the 30^th^ of April to the 30^th^ of May, the 3^rd^ of June, the 5^th^ of June, the 20^th^ of June, the 25^th^ of June to the 20^th^ of July, and the 23^rd^ of July to the 24^th^ of July were statistically significant.

The unadjusted OR for e-scooter injuries was 0.5 (CI 0.4-0.6) for the restricted period compared with the unrestricted period. When adjusted for hourly temperature (°C), rain amount (mm), wind speed (m/s), and visibility (km), the OR was 0.5 (CI 0.4-0.6).

There were fewer e-scooter injuries between 11 p.m. and 5 a.m. in the restricted period compared with the unrestricted period (Figure 2). Furthermore, the proportion of intoxicated drivers involved in accidents decreased from 38% to 31% (p<0.001) after introducing the restrictions. The reported proportion of helmet usage in the injured was 2% before restrictions and 6% after restrictions (p=0.08). The proportion of head injuries was 45% before the restrictions and 40% after the restrictions (p=0.11). The accident severity did not show a detectable difference between the study periods (p=0.49) (Table 1).

**Figure 2:**
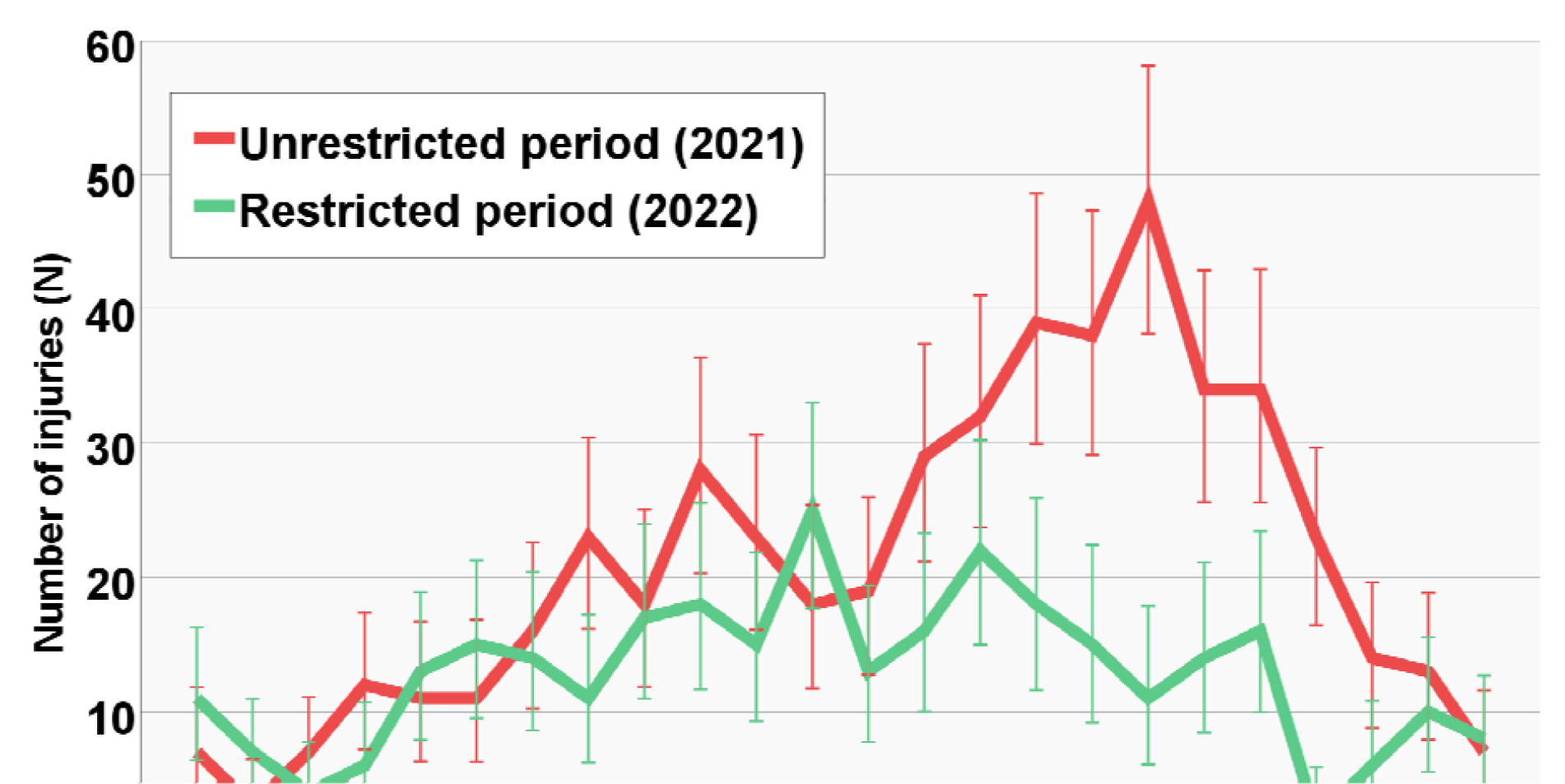
The number of e-scooter injuries at each time of the day. The vertical lines indicate 95% confidence intervals The restrictions afflicted: 1) Maximum speed limit 20km/h 2) Shared e-scooters were prohibited on Friday and Saturday nights between 12 p.m. and 5 a.m. 3) A nighttime speed limit of 15km/h for shared e-scooters between 12 p.m. and 5. a.m. from Sunday to Thursday The differences between 11 p.m. and 5 a.m. were statistically significant.

## Discussion

We found that the overall incidence of electric scooter injuries in Helsinki was distinctly lower after implementing restrictions that limited the top speed to 20km/h and restricted the availability of e-scooters at nighttime. The proportion of nighttime injuries and alcohol-related injuries also decreased. To our knowledge, this is the first study showcasing the efficacy of speed limitations and nighttime restrictions in preventing e-scooter injuries.

The top speed of the e-scooter seems to play an important role in the incidence of injuries. In Tampere, the third largest city in Finland, the incidence of e-scooter injuries was 18/100 000 trips from April 2019 to April 2021, which is practically equal to our study period with no restrictions (18). During the study period, Tampere had no specific e-scooter restrictions besides the 25 km/h speed limit. In 2022, the incidence of e-scooter injuries remained similar in Tampere (17/100 000 trips), even though in September 2021, the nighttime speed limit was lowered to 15 km/h in central areas, and since June 2022, the nighttime restrictions covered the whole city, but the daytime top speed was not lowered (19). In other countries, with no distinct restrictions on e-scooter usage, the rate of injuries is reported to be 20-21 injuries per 100 000 trips (20,21). On the contrary, Andersson and Djärv reported a lower incidence (3/100 000 trips) in Stockholm, Sweden, in 2019-2020, where the maximum speed limit for e-scooters is 20km/h. (22). Before the restrictions in Helsinki during the 2021 study period, the e-scooter-related injury incidence was 19/100 000, and after the restrictions, it decreased to 9/100 000. Lowering the maximum speed of e-scooters, even 5km/h, seems to decrease the injury burden substantially.

The primary aim of nighttime restrictions is to limit the number of intoxicated riders, and therefore limit injuries, as alcohol use has been common in patients suffering e-scooter injuries (23,24). To our knowledge, nighttime restrictions have only been studied in a single study conducted by Anderson et al. in the USA (25). Ultimately, they did not find evidence of a difference after the infliction of nighttime restrictions, although there were fewer accidents during the restriction period. In our study, the amount of both nighttime and alcohol-related injuries decreased after implementing the restrictions. We assume that nighttime restrictions are especially effective in populations with common evening and night-focused excessive alcohol usage. However, based on the literature discussed in the previous chapter, the limitation of overall top speed might be a more efficient method to decrease the overall injury burden.

The main uncertainty regarding our results is related to the possible bias caused by a learning curve regarding the use of e-scooters and the effect of increasing awareness related to the risks of these vehicles. Although it was impossible to adjust these phenomena, we consider that the effect of learning is minor compared to the restrictions. In Tampere, Finland, a city with a similar population and behavioural habits to Helsinki, the incidence of injuries remained fairly the same between 2019 and 2022, when there were no major restrictions (19). In addition, Williams et al. reported that the number of monthly rides and e-scooter-related emergency department visits correlated well, and there was no detectable decrease or increase in the monthly injury incidence during their study period from 8/2018 to 12/2019 (21). A study published by Farley et al. reported that the number of e-scooter injuries increased in the first three years after the introduction of shared e-scooters (11). Overall, these studies indicate that injury incidence tends not to decrease automatically during the long-term availability of e-scooters.

Other limitations of our study arise mainly due to its retrospective design. First, as we do not have a functional registry for e-scooter accidents, we had to rely on a word search from the hospital database. Therefore, not all accidents might have been collected. However, this limitation is similar during both study periods. Second, the intel regarding the ownership of the e-scooters was reported infrequently. Although we excluded injuries that were related to privately owned e-scooter according to the ED text, it is likely that our results still include an unknown proportion of injuries related to personally owned e-scooters. However, because privately owned e-scooters have also become more popular, it is unlikely that they would have caused a higher injury incidence in 2021 compared to 2022. Finally, some variables, for example, helmet use, were not routinely reported and cannot be compared reliably between the two periods.

## Conclusions

Even minimal restrictions on the top speed and nighttime usage of e-scooters seem to decrease the amount of hospital care requiring e-scooter injuries substantially. We recommend considering similar restrictions in cities where rental e-scooters are available.

## Data Availability

The data used in the study is available from a reasonable request

## List of abbreviations

E-scooter =: Stand-up electric scooter
ED =: Emergency department
AIS =: Abbreviated Injury Score
NISS =: New Injury Severity Score
IQR =: Inter Quartile Range

## Declarations

### Ethics approval and consent to participate

Organisational approval was gained from Helsinki University Hospital research board (HUS/44/2021). Based on Finnish legislation on medical research, utilising public and published data, registry and documentary data, and archive data do not require ethical board processing. Therefore, the research board exempted this study from requiring an ethical committee review and waived the need for informed consent to participate. We did not contact any patients, and all data was handled pseudonymised.

### Availability of data and materials

The data used in the study is available from a reasonable request.

### Transparency declaration

The lead author HV affirms that this manuscript is an honest, accurate, and transparent account of the study being reported. No important aspects of the study have been omitted and any discrepancies from the study as planned have been explained.

### Patient and Public Involvement Statement

The patients and public were not involved

### Authors’ contributions

HV planned and organised the study, data collection, and manuscript writing. OP and HV collected the data. OP and HV wrote the initial manuscript. HV analysed the data, and produced the figures. AK, MC and V-PH commented on the study protocol. All authors took part in the interpretation of the data.All authors revised, and approved the manuscript. HV acts as the guarantor for the work.

## Acknowledgements

We wish to thank the biostatistician Hanna Granroth-Wilding for the consultation services and Linda Toppari for the help with data collection. In addition, we wish to thank the city of Helsinki and the e-scooter operators for their co-operation in establishing the restrictions. We thank Helsinki Urban Environment Division for providing the e-scooter usage data.

